# White matter connectivity in resilience in a general population sample of 12,516 individuals

**DOI:** 10.1101/2023.03.06.23286759

**Authors:** Baukje S. de Vries, Nic J.A. van der Wee, Steven J.A. van der Werff

## Abstract

**Objective:** Neurobiological correlates of resilience to stress or trauma have not been investigated extensively. Studies on white matter connectivity in resilience, in particular, have been few and far between. This explorative study included 12,516 participants from the UK Biobank to investigate white matter connectivity in resilience in 13 selected regions of interest (ROIs).

**Methods:** The data for this cross-sectional cohort study were retrieved from UK Biobank, a large-scale biomedical database and research resource. The study included 40-69-year-old participants divided into three groups: a trauma-exposed, healthy (resilient) group, a trauma-exposed, mentally ill (vulnerable) group and a nonexposed, healthy (control) group. The primary outcome measures consisted of mean fractional anisotropy values in the following ROIs: cingulate gyrus, cingulum hippocampus, superior fronto-occipital fasciculus, uncinate fasciculus, corpus callosum body, genu, splenium, and tapetum. Group differences in fractional anisotropy were assessed using a one-way ANOVA.

**Results:** This study did not find correlates of resilience in the investigated ROIs.

**Conclusions:** The results indicate that there is no association between resilience and white matter connectivity in the selected ROIs in this large population sample, which is in contrast with previous studies on morphometric and functional correlates of resilience. To corroborate this finding, the authors recommend collection of longitudinal data and utilization of voxelwise analysis, if possible, to improve sensitivity. The authors also suggest a whole-brain analysis in a similar study population to investigate other white matter tracts that might be implicated in resilience.

## Introduction

In the preceding decades, researchers in the field of psychiatry and neuroscience have become increasingly interested in the neurobiology related to the concept of resilience. Resilience is often defined as a dynamic mechanism that consists of positive adaptation within the context of significant adversity, and furthermore, from a more psychobiological standpoint, as short- and long-term adaptive processes that reduce allostatic load. (1-4) In studies on resilience, it is often more plainly operationalized as the absence of psychopathology after experiencing a traumatic event. Studying resilience to trauma could contribute to the development of new treatment and prevention strategies for individuals that are at high risk of traumatic exposure. It is important to note that, while resilience has already been studied more extensively in its psychological and social context, neurobiological research on resilience, in particular research on neural mechanisms, is lagging behind. The main focus of neuroimaging of resilience so far has been on morphometric and functional correlates of resilience, using magnetic resonance imaging (MRI). (4) Contrastingly, white matter integrity in resilience has, to our knowledge, been studied to a limited extent and only in small sample sizes, even though the interconnectivity of brain regions through white matter tracts could very well be implicated in resilience.

So far, two animal studies have investigated white matter connectivity in resilience models. One study showed lower fractional anisotropy (FA), a measure of white matter integrity, in the hippocampus, hypothalamus, nucleus accumbens, cingulate cortex and amygdala for resilient (social stress exposed, normal behaving) mice compared to vulnerable (social stress exposed, socially avoidant) mice. (5) In contrast, another study, reported no significant FA differences between resilient, control (nonexposed, normal behaving) and vulnerable mice. (6)

In humans, studies on white matter integrity in resilience are also scarce and of heterogeneous designs. The available data consists mostly of studies focused on small samples of high-risk individuals such as first responders and military personnel. Studies typically only include patients with post-traumatic stress disorder (PTSD) in the group with mental illness and trauma exposure, even though a traumatic event is also known to be a precipitating factor for many other psychiatric disorders. (7-10) Importantly, the data is scarce to such a degree that researchers interested in white matter correlates of resilience have had to make do with studies that are focused on vulnerability.

These studies usually compare a healthy, trauma-exposed (resilient) group to a mentally ill, trauma-exposed (vulnerable group). This is far from ideal, since, without a healthy, nonexposed (control) group, it is not possible to distinguish the neural correlates of resilience and those of mental illness. (4)

Some studies have used a design with such an additional non-exposed healthy group. A study by O’Doherty et al. found lower FA in the uncinate fasciculus (UF), cingulum cingulate gyrus (CG), superior longitudinal fasciculus, corpus callosum body (CCB), genu (CCG) and splenium (CCS) in a trauma-exposed, healthy group and in a PTSD group compared to a healthy, non-exposed control group. (11) A second study by van der Werff et al. investigated white matter connectivity in highly resilient police officers with whole brain analysis and found an increase in structural connectivity in the corticopontine tract in the resilient group. (8) The aforementioned studies investigated samples of 75 and 81 people, respectively. A larger sized study by the PGC-ENIGMA PTSD consortium (N=3047), though not designed specifically to detect correlates of resilience, showed higher FA in the corpus callosum tapetum (CCT) and superior fronto-occipital fasciculus (SFOF) for the resilient trauma-exposed, non PTSD (TENP) group (N=113) compared to the trauma-exposed, PTSD group (N=2704) and lower FA in CCT, CCS and fornix or stria terminalis in the TENP group compared to nonexposed controls (N=180). (12) All in all, white matter tracts most likely to be implicated in resilience, as evidenced by what has been published so far on resilience and vulnerability, are the CG, cingulum hippocampus (CH), SFOF, UF, CCB, CCG, CCS and CCT. (4, 8, 11-20)

To investigate integrity of these white matter tracts in the context of resilience in a large population sample, we conducted a cross-sectional cohort study with diffusion tensor imaging (DTI) scans available from the UK Biobank, a large-scale biomedical database and research resource based in the United Kingdom. The resource, which includes lifestyle and health information, blood samples, heart and brain scans and genetic data of the 500,000 volunteer participants, is globally accessible to approved researchers who are undertaking health-related research that’s in the public interest.

The aim of this study was to investigate mean FA values in the CG, CH, SFOF, UF, CCB, CCG, CCS and CCT to determine white matter integrity in resilient brains compared to vulnerable and control brains. Due to the scarcity of the available evidence, this study was explorative in nature and therefore no hypothesis was defined.

## Methods

### Participants

We received approval from UK Biobank to conduct this study under project number 31102. All data and materials are available via UK Biobank. UK Biobank has approval from the North West Multi-centre Research Ethics Committee (MREC) as a Research Tissue Bank (RTB) approval. (21) The participants were drawn from the source population of the UK Biobank, which consists of 40-69-year-old residents of the United Kingdom who volunteered to participate. (22) Participants were requested to attend one or more assessments, including a general assessment that included several questionnaires, and an imaging assessment.

All participants were right-handed and had completed the imaging assessment and the questionnaire on mental health. We excluded patients that reported mental illnesses that are known to have a strong genetic component. We also excluded patients that reported a somatic diagnosis that would impact brain structure or connectivity. Thus, exclusion criteria were: past or current diagnosis of a psychotic disorder, bipolar disorder, obsessive compulsive disorder, any phobia except for agoraphobia, attention deficit or attention deficit and hyperactivity disorder or an autism spectrum disorder; diagnosis of any neurological injury, illness or a neurological developmental or chronic problem; diagnosis of an endocrinological problem that that could impact brain structure or connectivity; withdrawal from the UK Biobank study, as communicated through UK Biobank.

### Group selection

Subjects were divided into three groups. The groups included a resilient group (scoring positive on having experienced a traumatic event, and reporting no life-time mental illness), a vulnerable group (scoring positive on having experienced a traumatic event and reporting a diagnosis during their lifetime of one or more mental illnesses) and a control group (scoring negative on having experienced a traumatic event, and reporting no life-time mental illness). We excluded the group that scored negative on trauma exposure and positive on mental illness.

For assessment of trauma exposure, four binomial variables were used, in addition to three ordinal variables that used a five-point scale (never, rarely, sometimes, often, very often). Physical and sexual abuse as a child as well as domestic abuse by an (ex-)partner in adulthood were considered traumatic events if the participant reported to have experienced this sometimes, often or very often. Having been in a life-threatening accident, having been victim of physically violent crime, having witnessed sudden violent death or having been victim of sexual assault were also classified as traumatic events.

Mental illness, or a history thereof, was defined as having reported one or more mental health problems ever diagnosed by a professional or having ever been addicted to any substance (not including caffeine and tobacco) or behavior (such as gambling).

At this time, to make a clear distinction between the resilient and the control group, we excluded the control group participants that had rarely experienced physical or sexual abuse as a child or domestic abuse by an (ex-)partner in adulthood. Thus, the control group consisted of participants that had never experienced any of these events. For the same reason, we excluded controls that had rarely, sometimes, often or very often experienced sexual interference by (ex-)partner without consent, had felt hated by a family member as a child or had been belittled by an (ex-)partner as an adult. Lastly, we excluded participants from the control group if they had been involved in combat or if they had been diagnosed with a life-threatening illness.

### Questionnaires

Information derived from the questionnaires on mental health, medical conditions, early life factors and education was used for the inclusion and the assessment of education level. The baseline characteristics of sex and age were derived from the central registry of participants. The mental health questionnaire input was also used to classify a participant as either resilient, control or vulnerable. To determine highest attained level of education by the participants, their reported qualifications were divided into three categories. Basic education consisted of primary education or no school education at all, secondary education was classified as a high school education, and tertiary education was defined as having received one or more years of university education or having received a vocational degree.

At the UK Biobank assessment center, questionnaires were, whenever possible, administered using a touchscreen computer on which participants could work independently. For data fields that required detailed questioning, such as taking a medical history, computer-assisted personal interviews were held by members of staff. Participants were also requested to gather information on medical history beforehand to improve recall. (22)

### MRI procedures

A Siemens Skyra 3T scanner with a standard Siemens 32-channel RF receive head coil was used for the imaging assessments. The DTI images were acquired using a spin-echo echo-planar sequence with voxel dimensions of 2×2×2 mm. Five T2-weighted (b=0 s/mm^2^) baseline volumes, 3 blip-reversed T2-weighted (b=0 s/mm^2^) baseline volumes, 50 b=1000 s/mm^2^ and 50 b=2000 s/mm^2^ diffusion-weighted volumes were acquired with in total 100 distinct diffusion-encoding directions. (23)

A basic automated quality check was performed to remove unusable or corrupted data. The images were corrected for eddy currents and head motion, and outlier slices were corrected, using the Eddy tool. (24) The DTI outputs of FA, mean diffusivity (MD), axial diffusivity (AD) and radial diffusivity (RD) were created using the DTI fitting tool DTIFIT. Tract-based spatial statistics (TBSS) was used to create a study-specific FA skeleton using the standard-space tract masks from the research group at the Johns Hopkins University, which resulted in the predefined mean FA white matter tracts that were used for this study. (23, 25-27)

### Statistics

To compare baseline characteristics between groups, we used a chi-square test, a one-way ANOVA and a Kruskall-Wallis test for the categorical, continuous and ordinal variables, respectively. Group differences in white matter FA for each ROI were assessed using a one-way ANOVA. Additionally, in the event of significant differences between groups regarding sex or age, this characteristic was added as a covariate. If the difference from the mean for either of these baseline variables was more than 5% for any one of the groups, a mediation analysis was performed instead of adding the variable as a covariate.

If the ANOVA was significant, we conducted three one-tailed T tests comparing FA means between the three groups as our post hoc tests. If applicable, the covariate that had been added in the FA analysis was added in this analysis as well. In the event that such a post hoc T test between two groups proved significant, MD, RD and AD means for that region of interest were compared between the two groups to facilitate further interpretation. For this we used one-tailed T tests, seeing as the direction of the possible association could be derived from the FA results. To represent effect size, we used Cohen’s *d*, which we calculated using the effect size calculator by Lenhard and Lenhard. (28)

To control for multiple testing, we corrected the results of the statistical tests with the adjusted Bonferroni correction by Li and Ji. (12, 29). This method included a calculation of the effective number of independent tests (M_eff_) and took into account the assumption that the statistical tests were not independent, since the studied ROIs were interconnected white matter tracts. For the FA ROIs, the correlation coefficient ranged between 0.101 and 0.813. Using the eigenvalues of the correlation matrix, we calculated a M_eff_ of 9, which yielded a threshold for significance of 0.05/9=0.0057.

Based on the results, it was decided to do a post hoc analysis using a more narrow definition of trauma exposure, focusing on childhood trauma which was expected to have the biggest impact on connectivity. Two variations were used: in the first, wider definition, the responses “sometimes”, “often” and “very often” for physical or sexual abuse as a child were included, in the second, narrower definition, only “often” and “very often” were included. It was expected that using these definitions of more severe, early trauma, the resilient and vulnerable brains would show an altered pattern of connectivity.

## Results

We received mental health data and DTI data on 18589 participants from the UK Biobank, of which 16554 were right-handed. After exclusion of participants with neurological or endocrinological problems or psychiatric exclusion criteria, 14586 participants remained. Of this number, 11184 participants had not experienced a mental illness as defined by our criteria, of which a total of 3070 participants had experienced a traumatic event and thus were classified as resilient. After completion of exclusion criteria for controls in the remaining 8114 participants, 7577 controls were included in the study. Out of the base population of 14684, 3402 participants reported a history of mental illness. Of this number, 1771 participants had experienced a traumatic event and were thus classified as vulnerable.

Baseline characteristics for the resilient, control and vulnerable groups are displayed in Table 1. The proportion of women was significantly higher in the vulnerable group (61.7%) as compared to the resilient group (44.1%) and control group (50.2%, p<0.001). Furthermore, the vulnerable group was on average significantly younger (61.1 years of age) compared to the resilient group and the control group (63.0 and 63.2 years respectively, p<0.001). With regard to highest level of education, basic and secondary education were more prevalent in the control group and tertiary education was more prevalent in the resilient and vulnerable groups (p<0.001). Because of the differences in mean age, we added age to the analyses as a covariate. We also performed a mediation analysis for the effect of sex on the results of the analyses. We did not correct for level of education because we hypothesized that level of education, as an index of intelligence, would impact white matter connectivity. Studies on resilience and education have shown that resilient individuals more often pursue higher education, thus level of education would be in the causal pathway for the effect of resilience on white matter connectivity. (30)

**Table 1:** Baseline characteristics.

|  | Resilient<br>(N=3070) |  | Control<br>(N=7577) |  | Vulnerable<br>(N=1869) |  | Total<br>(N=12516) |  | *p* |
| --- | --- | --- | --- | --- | --- | --- | --- | --- | --- |
|  | *N* | % | *N* | % | *N* | % | *N* | % |  |
| Sex, female | 1355 | 44.1 | 3805 | 50.2 | 1093 | 61.7 | 6253 | 50.4 | <0.001 |
| Highest education level<sup>a</sup> |  |  |  |  |  |  |  |  | <0.001 |
| Basic | 299 | 10.0 | 991 | 13.3 | 206 | 11.9 | 1496 | 12.3 |  |
| Secondary | 784 | 26.1 | 2093 | 28.1 | 442 | 25.5 | 3319 | 27.3 |  |
| Tertiary | 1922 | 64.0 | 4352 | 58.5 | 1085 | 62.6 | 7359 | 60.4 |  |
|  | *Mean* | *SD* | *Mean* | *SD* | *Mean* | *SD* | *Mean* | *SD* |  |
| Age | 63.0 | 7.3 | 63.2 | 7.5 | 61.1 | 7.3 | 62.9 | 7.5 | <0.001 |

<sup>a</sup> Missing value in 2.0% of participants.

The most common trauma mechanisms in both the resilient and vulnerable group were: having been victim of a violent crime (40.1% and 38.2%, respectively), having witnessed sudden violent death (32.1% and 25.9%, respectively) and having been victim of sexual assault (26.4% and 41.0%, respectively, see ST1).

Comparing the FA of the resilient group, control group and vulnerable group, we found significant differences in the right CG (p=0.044, see Table 2) and also in the left and right SFOF (p=0.002 and p<0.001, respectively). After adjusted Bonferroni correction, the results for the SFOF left and right remained significant.

**Table 2:**
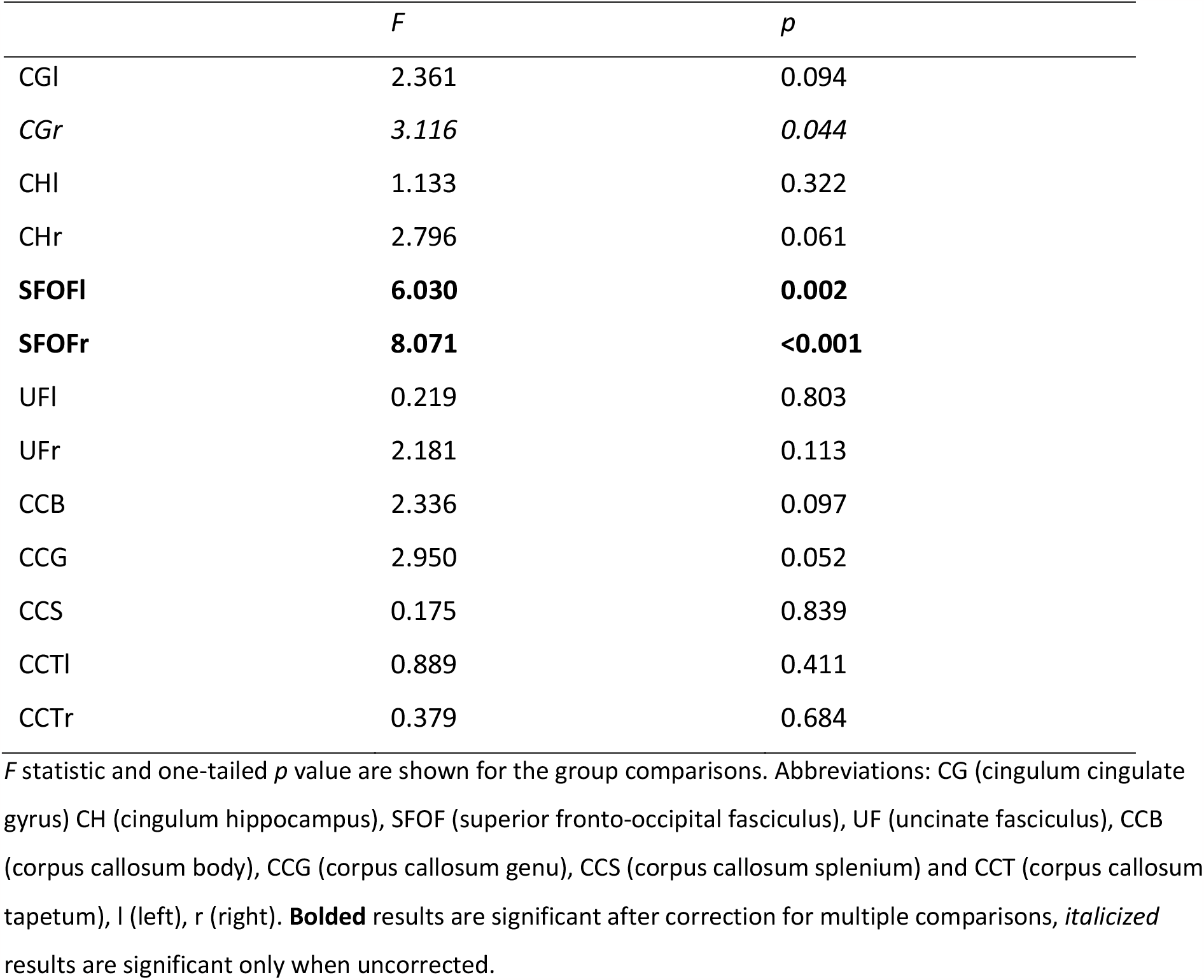
Results for the FA group comparisons using one-way ANOVA with age as a covariate.

The mediation analysis demonstrated an effect of sex on the results for the right CG: after correction for sex, the result did not remain significant (p=0.258). The significant results for the left and right SFOF survived correction for sex (p=0.003 and p=0.001 respectively).

One-tailed post hoc T tests were performed on the ROIs with significant results (see Table 3). The comparison between the resilient and the control group was not statistically significant in any of the ROIs. We found significantly higher FA in the left and significantly lower FA in the right SFOF in the vulnerable group compared to the control group and the resilient group. We also found significantly lower FA in the vulnerable group in the right CG. Effect sizes in the significant comparisons between the resilient and vulnerable group, expressed as Cohen’s *d*, ranged from 0.053 to 0.086. The largest effect sizes (*d*=0.080 and *d=*0.086) could be attributed to the left and right SFOF, respectively. For the comparisons between the vulnerable and control group, effect sizes ranged between 0.067 and 0.102. Similarly, the largest effect sizes could be attributed to the left and right SFOF (d=0.090 and d=0.102, respectively).

**Table 3:** Results for the post hoc T tests on FA for significant ROIs.

|  | Resilient group vs control group |  |  | Resilient group vs vulnerable group |  |  | Vulnerable group vs control group |  |  |
| --- | --- | --- | --- | --- | --- | --- | --- | --- | --- |
|  | *t* | *d* | *p* | *t* | *d* | *p* | *t* | *d* | *p* |
| CGr | 0.333 | 0.007 | 0.370 | 1.776 | 0.053 | 0.038 | -2.542 | 0.067 | 0.006 |
| SFOFI | -0.663 | 0.014 | 0.254 | -2.673 | 0.080 | 0.004 | 3.411 | 0.090 | <0.001 |
| SFOFr | -1.230 | 0.026 | 0.109 | 2.893 | 0.086 | 0.002 | -3.874 | 0.102 | <0.001 |

*t* statistic, Cohen's *d* and one-tailed *p* value are shown for the group comparisons. In the comparison of group A vs group B, a negative *t* statistic indicates lower FA in group A. Abbreviations: CG (cingulum cingulate gyrus), SFOF (superior fronto-occipital fasciculus), l (left), r (right). Significant results are *italicized*.

To compare the resilient group with the vulnerable group and the vulnerable group with the control group on AD, RD and MD, two tailed independent T tests including the covariate age were performed (see Table 4). For RD and MD in the right CG, significant differences were found between the resilient and vulnerable group (p=0.008 and p=0.011, respectively). For RD, a difference was found between the vulnerable and the control group (p=0.034). Comparing the resilient group to the vulnerable group on AD, RD and MD in the left SFOF, all differed significantly (p=0.019, p=0.004, p=0.004, respectively). Comparing the vulnerable group to the control group in the left SFOF, RD and MD showed a significant difference (p=0.003 and p=0.007, respectively). In the right SFOF, there were significant differences between the resilient and vulnerable groups on RD and MD (p=0.002 and p=0.008, respectively). In the right SFOF, there was also a significant difference on RD and MD between the vulnerable group and the control group (p=0.005 and p=0.045, respectively).

**Table 4:** Mean values and T test comparisons with age as a covariate for the AD, RD and MD for the resilient group vs the vulnerable group and for the vulnerable group vs the control group.

|  | Mean values |  |  | T test comparison |  |  |  |
| --- | --- | --- | --- | --- | --- | --- | --- |
|  | Resilient | Vulnerable | Control | Resilient group vs vulnerable group |  | Vulnerable group vs control group |  |
|  | group | group | group | *t* | *p* | *t* | *p* |
| CGr |  |  |  |  |  |  |  |
| AD | 1.329·10<sup>-3</sup> | 1.330·10<sup>-3</sup> | 1.332·10<sup>-3</sup> | -0.676 | 0.250 | -1.617 | 0.053 |
| RD | 4.674·10<sup>-4</sup> | 4.679·10<sup>-4</sup> | 4.687·10<sup>-4</sup> | -2.398 | *0.008* | -1.833 | *0.034* |
| MD | 7.546·10<sup>-4</sup> | 7.553·10<sup>-4</sup> | 7.565·10<sup>-4</sup> | -2.289 | *0.011* | -0.346 | 0.365 |
| SFOFI |  |  |  |  |  |  |  |
| AD | 1.164·10<sup>-3</sup> | 1.161·10<sup>-3</sup> | 1.167·10<sup>-3</sup> | *2.086* | *0.019* | -1.475 | 0.070 |
| RD | 5.434·10<sup>-4</sup> | 5.404·10<sup>-4</sup> | 5.447·10<sup>-4</sup> | *2.686* | *0.004* | -2.737 | *0.003* |
| MD | 7.504·10<sup>-4</sup> | 7.473·10<sup>-4</sup> | 7.522·10<sup>-4</sup> | *2.644* | *0.004* | -2.450 | *0.007* |
| SFOFr |  |  |  |  |  |  |  |
| AD | 1.157·10<sup>-3</sup> | 1.153·10<sup>-3</sup> | 1.160·10<sup>-3</sup> | 1.064 | 0.144 | -0.300 | 0.382 |
| RD | 5.336·10<sup>-4</sup> | 5.314·10<sup>-4</sup> | 5.348·10<sup>-4</sup> | *2.856* | *0.002* | -2.582 | *0.005* |
| MD | 7.413·10<sup>-4</sup> | 7.385·10<sup>-4</sup> | 7.433·10<sup>-4</sup> | *2.442* | *0.008* | -1.700 | *0.045* |

Mean values are shown for the three groups, *t* statistic and one-tailed *p* value are shown for the group comparisons. Abbreviations: CG (cingulum cingulate gyrus), SFOF (superior fronto-occipital fasciculus), l (left), r (right), AD (axial diffusivity), RD (radial diffusivity), MD (mean diffusivity). Significant results are *italicized*.

Finally, in the post hoc analyses on childhood maltreatment, the resilient and vulnerable groups consisted of 625 and 525, respectively, in the wider definition of childhood maltreatment, and of 85 and 121, respectively, in the narrower definition. In both analyses, age was added as a covariate to the analyses. In the first analysis, the one-way ANOVA was significant in the CGl, CGr, CHl, SFOFl, SFOFr, UFr and CCG and the results for the SFOFl and SFOFr survived correction for multiple testing. The post hoc T tests did not demonstrate a significant difference between the resilient and the control group in any of the ROIs. In the second analysis (with the narrowest definition), the one-way ANOVA showed significant differences in the CGr, CHr and CCG, but only when not corrected for multiple testing. Explorative post hoc T tests for this result showed a significant difference in the CGr and CHr between the resilient and control group (p=0.048 and p=0.003, respectively) as well as between the resilient and vulnerable group (p=0.007 and p=0.008, respectively).

## Discussion

This study sought to identify white matter correlates of resilience in a large population sample of 40-69-year-old volunteers from the UK Biobank. We compared three groups of subjects: a trauma-exposed, healthy (resilient) group, a trauma-exposed, mentally ill (vulnerable) group and a nonexposed, healthy (control) group. We did not find white matter correlates of resilience in the main analyses and also not in our post-hoc sensitivity analysis in childhood maltreatment subgroups.

There are several possible explanations for the absence of a clear association of white matter connectivity with resilience in the selected ROIs. First of all, since mean tract values were used we could have missed an effect that only voxelwise analyses would have detected. Secondly, the UK Biobank study population is reflective of a general, middle-aged population, which is in contrast with the populations of carefully selected and well-trained professionals such as special forces or first-responders typically included in most studies on resilience so far. In our case, opting for a broader population sample also necessitated expanding the definition of trauma that was used. Including, for example, domestic abuse, as well as experiencing a life-threatening accident and witnessing violent death as traumatic events, resulted in the inclusion of a wide variety of traumatic experiences, as shown in Table 5. As some studies suggest that different traumatic experiences could each have distinctive biological signatures, one may argue that lumping these various traumatic experiences might have reduced the power. Our post-hoc analysis in a population with a more homogeneous definition of trauma, however, also revealed no specific correlates of resilience to more severe forms of childhood maltreatment in subjects from this general population sample.

**Table 5:** Trauma mechanisms for the resilient and vulnerable group.

|  | Resilient (N=3070) |  | Vulnerable (N=1771) |  | Total (N=4841) |  |
| --- | --- | --- | --- | --- | --- | --- |
|  | *N* | % | *N* | % | *N* | % |
| Physical child abuse | 475 | 15.5 | 377 | 21.3 | 852 | 17.6 |
| Sexual child abuse | 199 | 6.5 | 200 | 11.3 | 399 | 8.2 |
| Physical abuse in adulthood | 294 | 9.6 | 323 | 18.2 | 617 | 12.7 |
| Life-threatening accident | 604 | 19.7 | 305 | 17.2 | 909 | 18.8 |
| Victim of violent crime | 1230 | 40.1 | 677 | 38.2 | 1907 | 39.4 |
| Witness of violent death | 984 | 32.1 | 459 | 25.9 | 1443 | 29.8 |
| Sexual assault | 811 | 26.4 | 726 | 41.0 | 1537 | 31.7 |

Most importantly, especially regarding the large sample size of this study, our results indicate that the regions of interest of this study are not correlated with resilience in the general, middle-aged population.

We found abnormalities in our vulnerable group as compared to the control and resilient group. There was increased mean FA in the left and decreased mean FA in the right SFOF ROI. Higher FA is associated with greater white matter integrity, specifically, greater axon diameter, density or myelination. (31) Our findings for the left and right SFOF, a tract associated with attention and vigilance, may represent a vulnerability marker or a consequence of the various sequelae of the exposure to a traumatic event. (32, 33) Studies by Aschbacher et al. and the ENIGMA PTSD study by Dennis contradict each other on the results for the SFOF (12, 14) While also taking into account that both studies are quite different from our study with regard to design and inclusion criteria and longitudinal data are lacking, our findings for the SFOF are not easy to interpret. Moreover, from our results, it seems that the effect of vulnerability on MD and RD differs between white matter tracts. More investigation is required to interpret the combinations of increased and decreased MD and RD in the context of resilience and vulnerability.

We believe that the present study has several strengths. It is the largest to date to investigate white matter correlates of resilience. We were also able to investigate the effect of age and sex in relation to our outcome variables. In comparison to most other studies, we included a healthy, nonexposed control group and expanded our definition of trauma related psychopathology to include mental disorders other than PTSD, in particular affective disorders, to obtain a more accurate picture of vulnerability and resilience. Finally, the utilization of the Ji and Li adapted Bonferroni correction ensured correction for multiple testing tailored to our intercorrelated outcome variables.

There are some limitations to report as well. Ultimately, the UK Biobank database was not specifically designed for research into resilience. The retrospective nature of the UK Biobank questionnaires we used for the purpose of in-and exclusion and group selection introduced a risk of recall bias.

Furthermore, in the absence of longitudinal data, our study was not designed to make causal inferences, nor could it deduce whether white matter differences were the result of an acquired resilience or vulnerability, or if such differences were present in the participants prior to trauma exposure. As mentioned previously, the utilization of mean tract values instead of voxelwise data might have obscured an association with resilience. Out of necessity, our ROI selection was based on limited literature. Possibly, other white matter tracts could be implicated in resilience. Lastly, only white matter tracts were investigated in this study and it should be noted that previous studies have identified correlates of resilience using other morphometric and functional MRI approaches. (4)

In conclusion, our results could suggest that there are no specific white correlates of resilience in the general population. Clearly, this should be corroborated in future studies which should ideally focus on collecting longitudinal data as well as using validated questionnaires and patient records to minimize risk of recall bias. We also recommend that future studies on white matter correlates of resilience conduct voxelwise analyses if possible, to improve sensitivity, and investigate other white matter tracts that might be implicated in resilience.

## Data Availability

All data produced in the present study are contained in the manuscript

## Acknowledgement

This research has been conducted using data from UK Biobank, a major biomedical database. More information is available from www.ukbiobank.ac.uk.

## References

1. Charney DS. Psychobiological mechanisms of resilience and vulnerability: implications for successful adaptation to extreme stress. Am J Psychiatry. 2004;161(2):195–216.

2. Curtis WJ, Cicchetti D. Moving research on resilience into the 21st century: theoretical and methodological considerations in examining the biological contributors to resilience. Dev Psychopathol. 2003;15(3):773–810.

3. Cicchetti D, Rogosch FA. Adaptive coping under conditions of extreme stress: Multilevel influences on the determinants of resilience in maltreated children. New Dir Child Adolesc Dev. 2009;2009(124):47–59.

4. van der Werff SJ, van den Berg SM, Pannekoek JN, Elzinga BM, van der Wee NJ. Neuroimaging resilience to stress: a review. Front Behav Neurosci. 2013;7:39.

5. Anacker C, Scholz J, O’Donnell KJ, Allemang-Grand R, Diorio J, Bagot RC, et al. Neuroanatomic Differences Associated With Stress Susceptibility and Resilience. Biol Psychiatry. 2016;79(10):840–9.

6. Liu X, Yuan J, Guang Y, Wang X, Feng Z. Longitudinal in vivo Diffusion Tensor Imaging Detects Differential Microstructural Alterations in the Hippocampus of Chronic Social Defeat Stress-Susceptible and Resilient Mice. Front Neurosci. 2018;12:613.

7. Dohrenwend BP. Adversity, stress and psychopatholgy. New York: Oxford University Press; 1998.

8. van der Werff SJA, Elzinga BM, Smit AS, van der Wee NJA. Structural brain correlates of resilience to traumatic stress in Dutch police officers. Psychoneuroendocrinology. 2017;85:172–8.

9. Koenigs M, Huey ED, Raymont V, Cheon B, Solomon J, Wassermann EM, et al. Focal brain damage protects against post-traumatic stress disorder in combat veterans. Nat Neurosci. 2008;11(2):232–7.

10. Kuo JR, Kaloupek DG, Woodward SH. Amygdala volume in combat-exposed veterans with and without posttraumatic stress disorder: a cross-sectional study. Arch Gen Psychiatry. 2012;69(10):1080–6.

11. O’Doherty DCM, Ryder W, Paquola C, Tickell A, Chan C, Hermens DF, et al. White matter integrity alterations in post-traumatic stress disorder. Hum Brain Mapp. 2018;39(3):1327–38.

12. Dennis EL, Disner SG, Fani N, Salminen LE, Logue M, Clarke EK, et al. Altered white matter microstructural organization in posttraumatic stress disorder across 3047 adults: results from the PGC-ENIGMA PTSD consortium. Mol Psychiatry. 2019.

13. Meng L, Chen Y, Xu X, Chen T, Lui S, Huang X, et al. The neurobiology of brain recovery from traumatic stress: A longitudinal DTI study. J Affect Disord. 2018;225:577–84.

14. Aschbacher K, Mellon SH, Wolkowitz OM, Henn-Haase C, Yehuda R, Flory JD, et al. Posttraumatic stress disorder, symptoms, and white matter abnormalities among combat-exposed veterans. Brain Imaging Behav. 2018;12(4):989–99.

15. Li L, Lei D, Li L, Huang X, Suo X, Xiao F, et al. White Matter Abnormalities in Post-traumatic Stress Disorder Following a Specific Traumatic Event. EBioMedicine. 2016;4:176–83.

16. Fani N, King TZ, Jovanovic T, Glover EM, Bradley B, Choi K, et al. White matter integrity in highly traumatized adults with and without post-traumatic stress disorder. Neuropsychopharmacology. 2012;37(12):2740–6.

17. Abe O, Yamasue H, Kasai K, Yamada H, Aoki S, Iwanami A, et al. Voxel-based diffusion tensor analysis reveals aberrant anterior cingulum integrity in posttraumatic stress disorder due to terrorism. Psychiatry Res. 2006;146(3):231–42.

18. Wang HH, Zhang ZJ, Tan QR, Yin H, Chen YC, Wang HN, et al. Psychopathological, biological, and neuroimaging characterization of posttraumatic stress disorder in survivors of a severe coalmining disaster in China. J Psychiatr Res. 2010;44(6):385–92.

19. Koch SBJ, van Zuiden M, Nawijn L, Frijling JL, Veltman DJ, Olff M. Decreased uncinate fasciculus tract integrity in male and female patients with PTSD: a diffusion tensor imaging study. J Psychiatry Neurosci. 2017;42(5):331–42.

20. Kim SJ, Jeong DU, Sim ME, Bae SC, Chung A, Kim MJ, et al. Asymmetrically altered integrity of cingulum bundle in posttraumatic stress disorder. Neuropsychobiology. 2006;54(2):120–5.

21. UK Biobank. UK Biobank - Improving the health of future generations [Available from: http://www.ukbiobank.ac.uk [Accessed 10th September 2020].

22. Sudlow C, Gallacher J, Allen N, Beral V, Burton P, Danesh J, et al. UK biobank: an open access resource for identifying the causes of a wide range of complex diseases of middle and old age. PLoS Med. 2015;12(3):e1001779.

23. Alfaro-Almagro F, Jenkinson M, Bangerter NK, Andersson JLR, Griffanti L, Douaud G, et al. Image processing and Quality Control for the first 10,000 brain imaging datasets from UK Biobank. Neuroimage. 2018;166:400–24.

24. Andersson JLR, Sotiropoulos SN. An integrated approach to correction for off-resonance effects and subject movement in diffusion MR imaging. Neuroimage. 2016;125:1063–78.

25. Mori S, Wakana S, Zijl PCV, Nagae-Poetscher L. MRI atlas of human white matter: Am. Soc. Neuroradiol.; 2005.

26. Wakana S, Caprihan A, Panzenboeck MM, Fallon JH, Perry M, Gollub RL, et al. Reproducibility of quantitative tractography methods applied to cerebral white matter. Neuroimage. 2007;36(3):630–44.

27. Smith SM, Jenkinson M, Johansen-Berg H, Rueckert D, Nichols TE, Mackay CE, et al. Tract-based spatial statistics: voxelwise analysis of multi-subject diffusion data. Neuroimage. 2006;31(4):1487–505.

28. Lenhard W, Lenhard A. Calculation of effect sizes. [Available from: https://www.psychometrica.de/effect_size.html [Accessed 30th October 2020].

29. Li J, Ji L. Adjusting multiple testing in multilocus analyses using the eigenvalues of a correlation matrix. Heredity (Edinb). 2005;95(3):221–7.

30. Werner EE, Smith RS. Overcoming the odds. High risk children from birth to adulthood. Ithaca and London: Cornell University Press; 1992.

31. Beaulieu C. The basis of anisotropic water diffusion in the nervous system - a technical review. NMR Biomed. 2002;15(7-8):435–55.

32. Milner AD, Goodale MA. Two visual systems re-viewed. Neuropsychologia. 2008;46(3):774–85.

33. Wakana S, Jiang H, Nagae-Poetscher LM, van Zijl PC, Mori S. Fiber tract-based atlas of human white matter anatomy. Radiology. 2004;230(1):77–87.

